# A Predictive Nomogram for In-ICU Deterioration of Stage 1 Pressure Injuries: A Retrospective Study

**DOI:** 10.64898/2026.01.30.26345188

**Authors:** Chenyan Zhang, Wei Wang, Huina Xu

**Author notes:** Correspondence: Huina Xu. These authors contributed equally to this work (Chenyan Zhang, Wei Wang).

## Abstract

**Background:** Preventing Stage 1 pressure injuries (PIs) from worsening in the ICU is a key clinical challenge. Early prediction of high-risk patients enables targeted prevention. We aimed to develop a model for this progression using admission data.

**Methods:** In this retrospective cohort study, eligible ICU patients with Stage 1 pressure injuries were randomly allocated into training (70%) and validation (30%) sets. Predictors were selected using LASSO regression. A multivariable logistic regression model was constructed and visualized as a nomogram. Model performance was evaluated by discrimination (AUC), calibration, and clinical utility (decision curve analysis).

**Results:** A total of 278 patients were randomly divided into training (n=195) and validation (n=83) sets. LASSO regression identified four independent predictors: diabetes (OR: 3.266; 95% CI: 1.451–7.352), maximum norepinephrine dose (OR: 13.032; 95% CI: 1.212–140.137), use of pneumatic compression pumps (OR: 3.308; 95% CI: 1.444–7.579), and albumin level at ICU admission (OR: 0.836 per unit increase; 95% CI: 0.777–0.900). The nomogram demonstrated excellent discrimination, with an AUC of 0.810 (95% CI: 0.748–0.872) in the training set and 0.805 (95% CI: 0.696–0.914) in the validation set. Good calibration and clinical utility were confirmed.

**Conclusions:** A nomogram incorporating four readily available factors at ICU admission effectively predicts the risk of Stage 1 PI progression. This tool may aid early risk stratification and guide precise preventive measures.

## Introduction

Pressure injuries (PIs) remain a prevalent complication in ICU patients, contributing to extended hospitalization and increased healthcare costs [1,2]. Despite established prevention protocols, the incidence of PIs in intensive care settings persists at concerning levels [3]. Stage 1 PIs, presenting as non-blanchable erythema, indicate early tissue compromise but do not invariably progress to more severe, full-thickness wounds (⩾Stage 2) [4]. This variability underscores a key clinical dilemma: reliably identifying which patients with Stage 1 PIs will deteriorate rapidly remains challenging.

Current risk assessment tools show limited accuracy in predicting PI progression within the ICU [5]. This limitation contributes to suboptimal prevention strategies [6]. A predictive model using admission data could enable early risk stratification [7], allowing timely intervention before irreversible damage occurs [8]. While such models have been applied to other outcomes in critical care [9], a specific tool for Stage 1 PI progression remains unavailable [10].

Therefore, the primary objective of this study was to develop and validate a novel prediction model, visualized as a nomogram, to individually estimate the risk of progression from Stage 1 to severe PIs (⩾ Stage 2) in ICU patients using routinely collected admission data.

## Materials and methods

### Study population

This single-center, retrospective cohort study was conducted at The Affiliated Lihuili Hospital of Ningbo University. The detailed patient screening process is shown in Figure 1. Patients were included if they met the following criteria: (1) adult patients (≥18 years) admitted to the ICU between January 2022 and December 2025; (2) presence of at least one Stage 1 pressure injury (PI) identified within 24 hours of ICU admission; (3) first ICU admission during the current hospitalization. The diagnosis and staging of PIs were performed according to the latest international guidelines [11]. The exclusion criteria were as follows: (1) patients with Stage 2 or higher PIs at ICU admission; (2) patients with an ICU length of stay less than 48 hours; (3) patients who were readmitted to the ICU during the same hospitalization; (4) patients with major missing clinical data (>20% of key variables). This study was approved by the Medical Ethics Committee of The Affiliated Lihuili Hospital of Ningbo University (Approval No. IIT2026SL0013). The requirement for informed consent was waived due to the retrospective nature of the study. All procedures were conducted in accordance with the Declaration of Helsinki.

**Figure 1.**
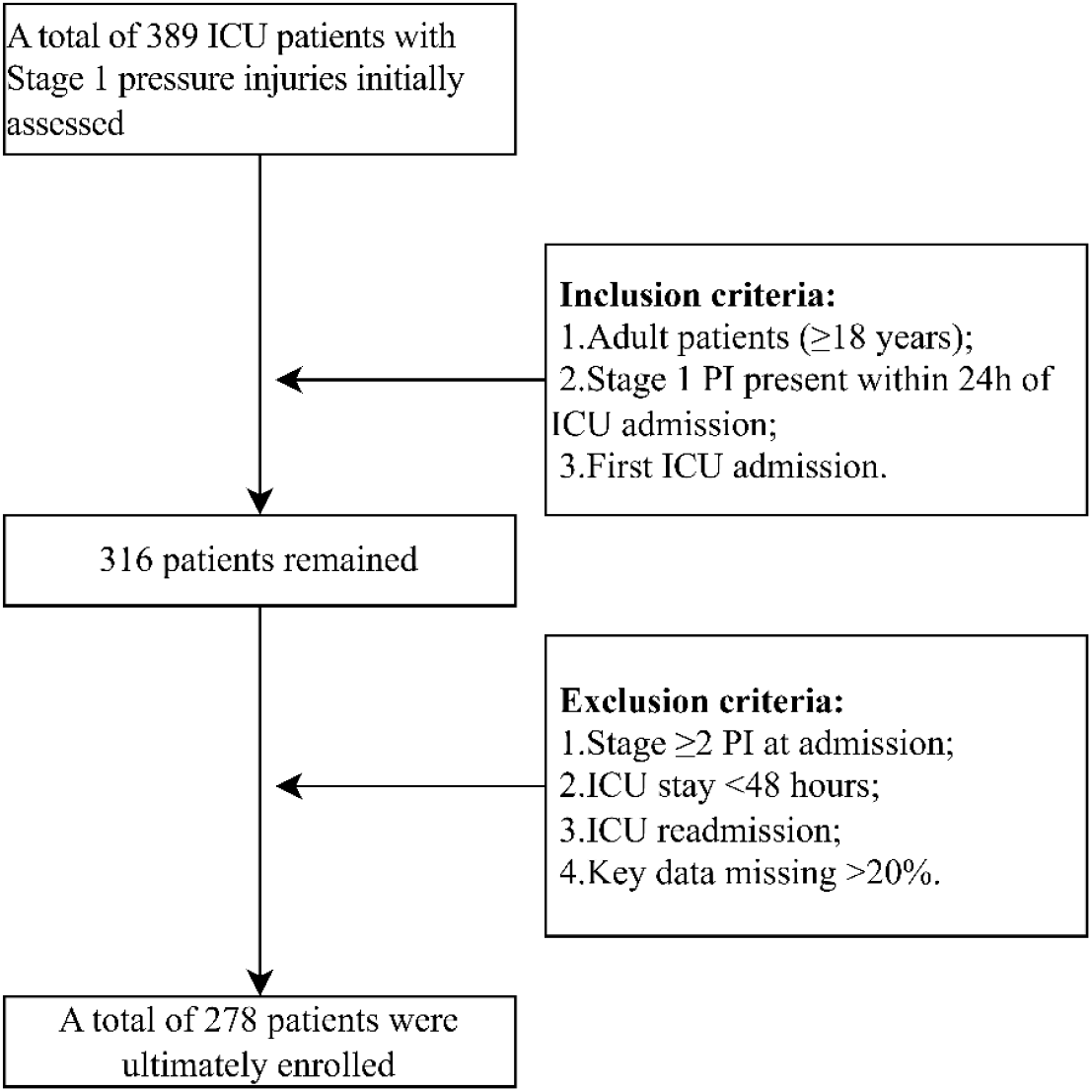
Flowchart of participant screening and enrollment in this cohort study; PI, pressure injury; ICU, intensive care unit

### Grouping and Data Collection

This study aimed to develop a prediction model for the progression of Stage 1 pressure injuries (PIs). Therefore, the study population was categorized into two groups based on the primary outcome: the progression group (patients whose Stage 1 PIs worsened to Stage 2 or beyond during their ICU stay) and the non-progression group (patients whose Stage 1 PIs did not worsen). This grouping allowed for a direct comparison of factors associated with the key clinical event. Data pertaining to demographic characteristics, disease severity scores, laboratory parameters upon ICU admission, and details of treatment interventions were retrospectively extracted from the hospital’s electronic medical record system. To ensure the model’s utility for early risk stratification, data collection was strictly focused on variables that are routinely available within the first 24 hours of ICU admission, such as APACHE II score, serum albumin level, and the use of critical care supports like mechanical ventilation and vasoactive drugs.

### Statistical analysis

Statistical analyses were performed using SPSS (version 22.0) and R software (version 4.2.0). Continuous variables, which did not conform to a normal distribution, were presented as medians with interquartile ranges (IQRs) and compared using the Mann–Whitney U-test. Categorical variables were expressed as counts and percentages, with between-group comparisons made using the chi-square test. The entire cohort was randomly divided into a training set and a validation set at a ratio of 7:3. Independent predictors were selected through Least Absolute Shrinkage and Selection Operator (LASSO) regression. A multivariable logistic regression model was subsequently constructed, and a nomogram was developed using the “rms” package in R. The predictive performance of the nomogram was evaluated by its discrimination (assessed using the receiver operating characteristic curve and quantified by the area under the curve), calibration (assessed using calibration curves), and clinical utility (assessed using decision curve analysis to estimate the net benefit across a range of threshold probabilities). A two-sided p-value < 0.05 was considered statistically significant.

## Results

### Patient Characteristics and Univariate Analysis

A total of 278 eligible ICU patients with Stage 1 PIs were enrolled in this study. The cohort was randomly divided into a training set (n=195, 70%) and a validation set (n=83, 30%). As detailed in Table 1, no statistically significant differences (all P> 0.05) were observed between the training and validation sets across all baseline characteristics, including demographics, clinical severity scores, comorbidities, interventions, and laboratory parameters. This indicates a successful and balanced randomization, ensuring the comparability of the two datasets for subsequent model development and validation.

**Table 1.**
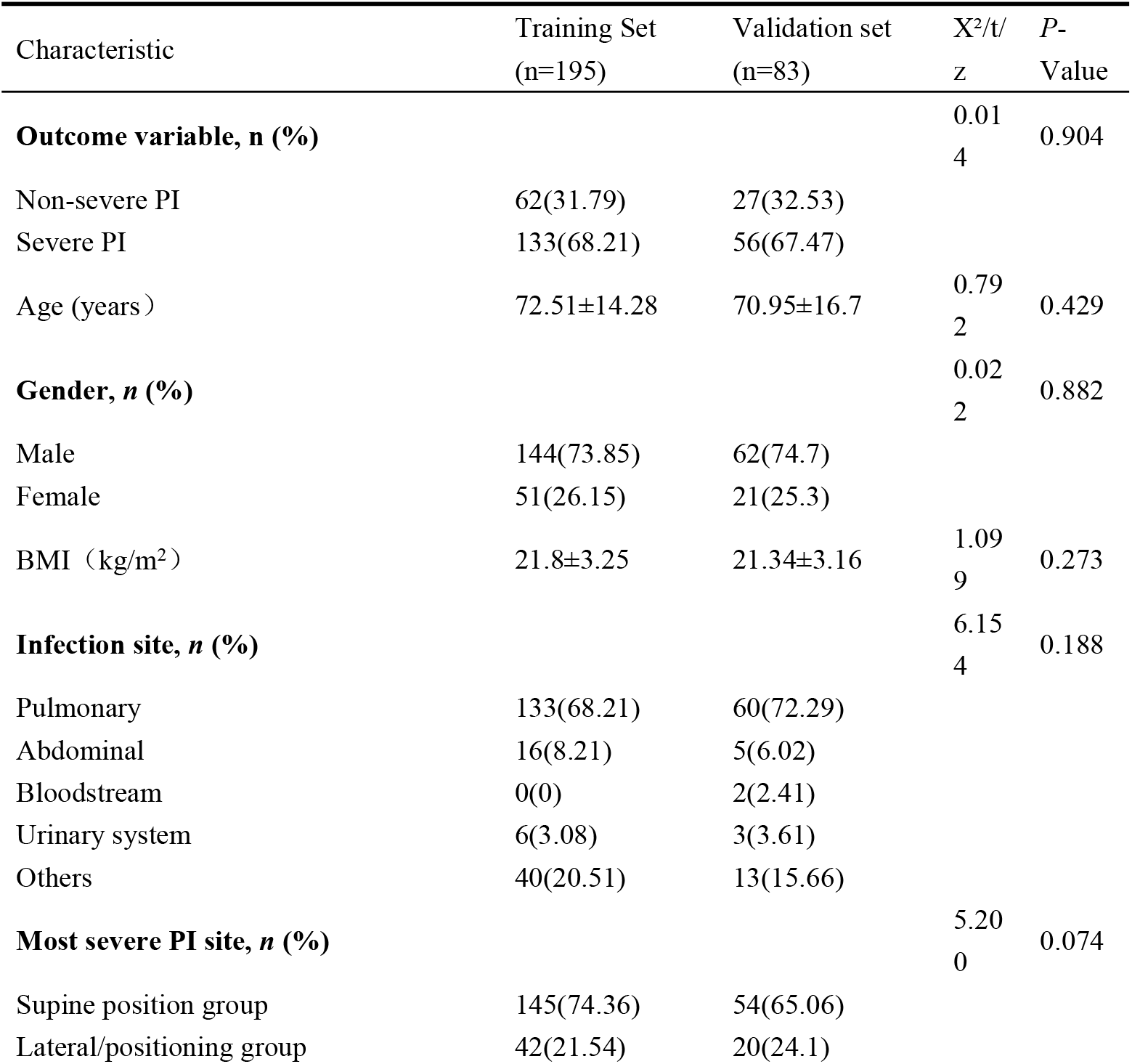
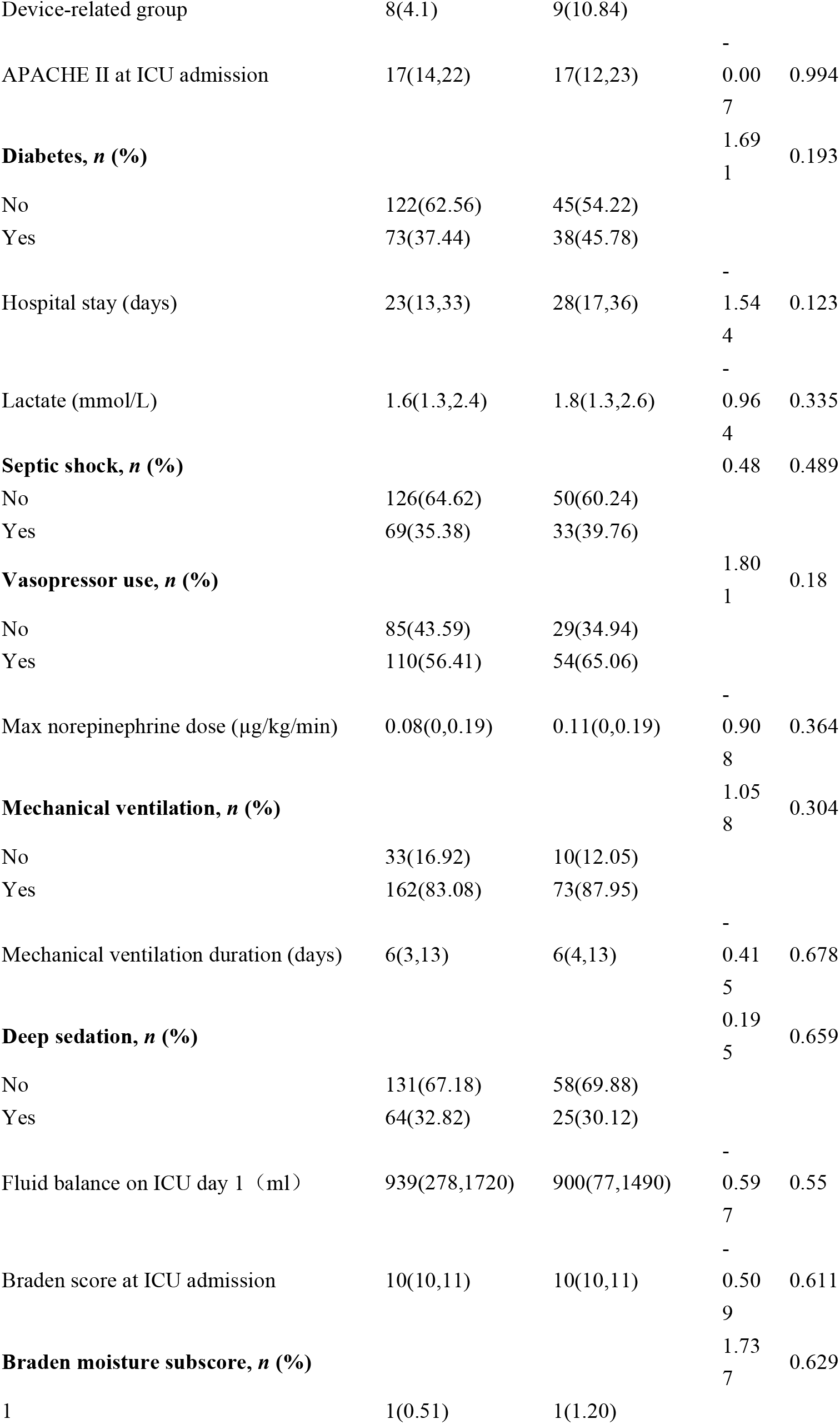
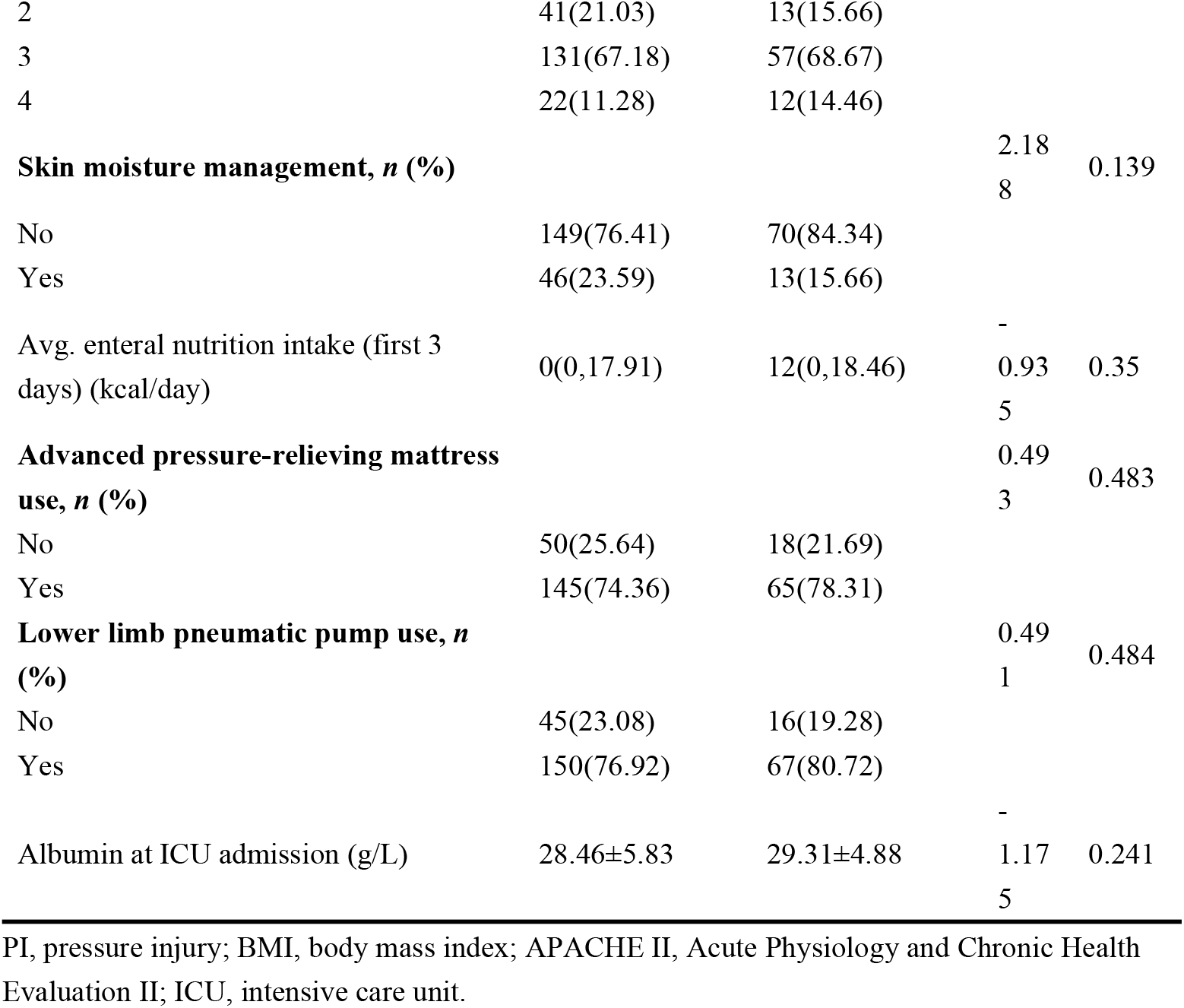
Comparison of baseline characteristics between the training and validation sets.

Table 2 presents a comparison of baseline characteristics between patients with non-severe and severe pressure injuries (PIs) in the training set. Significant differences were observed between the two groups. The severe PI group had a higher prevalence of diabetes (43.61% vs. 24.19%, p=0.009), required a greater maximum norepinephrine dose (median 0.08 vs. 0 µg/kg/min, p=0.047), had higher utilization rates of both advanced pressure-relieving mattresses (78.95% vs. 64.52%, p=0.032) and lower limb pneumatic pumps (82.71% vs. 64.52%, p=0.005), and exhibited a significantly lower serum albumin level at ICU admission (26.89 g/L vs. 31.81 g/L, p<0.001). No statistically significant differences were found in other demographic, clinical, or laboratory parameters.

**Table 2.**
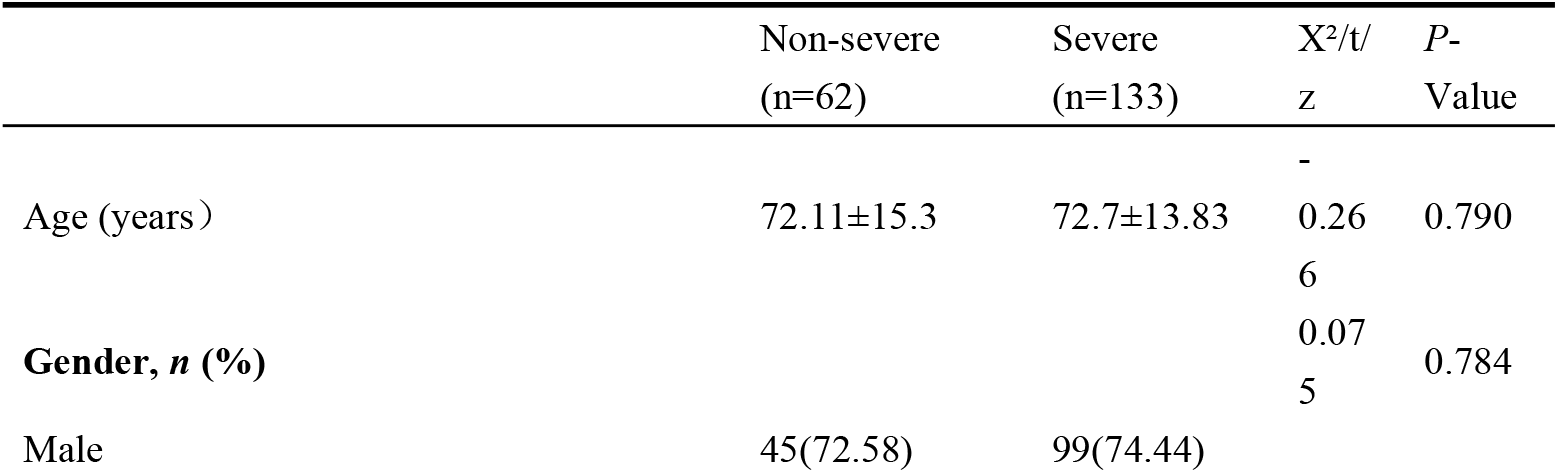
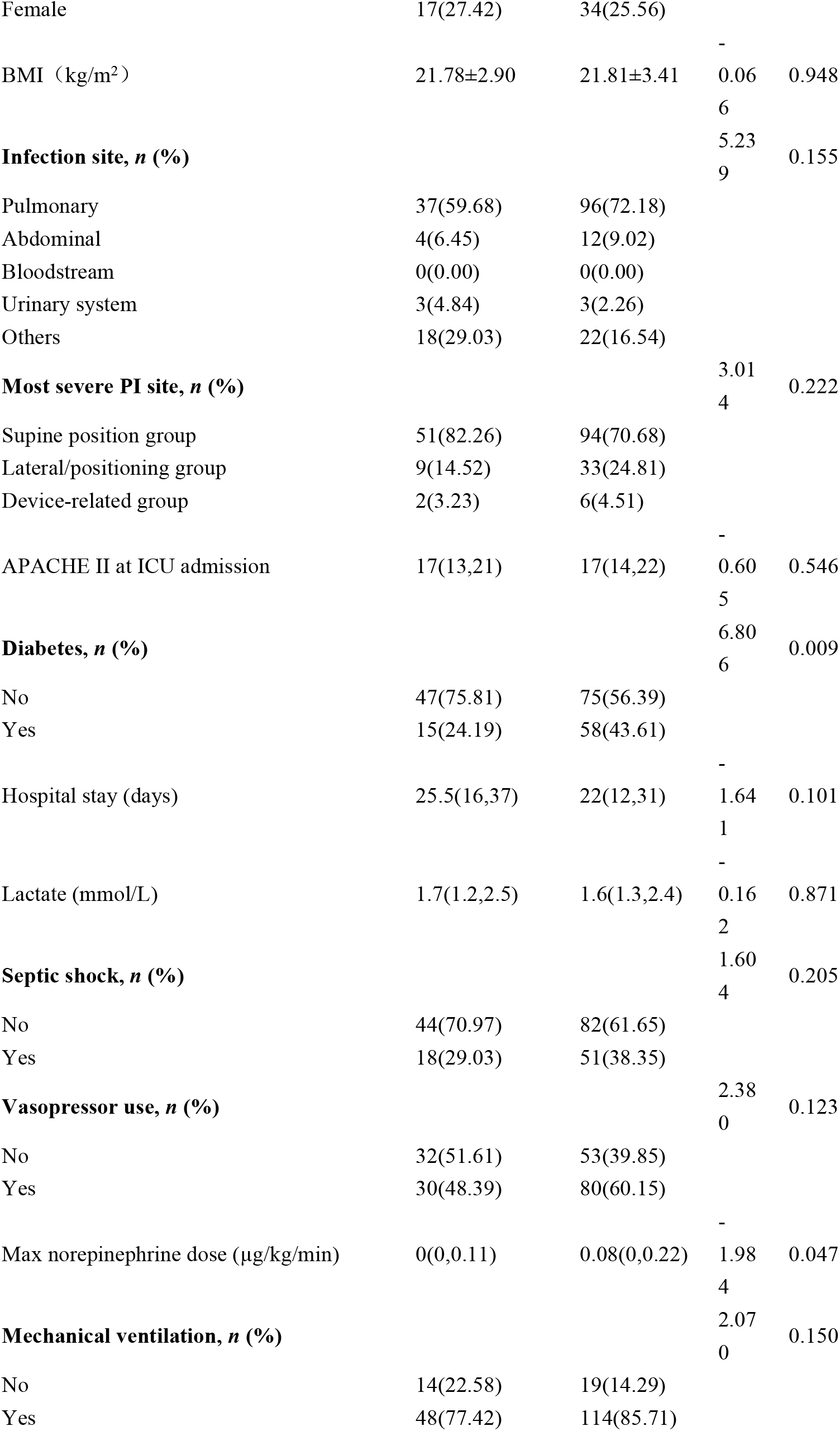
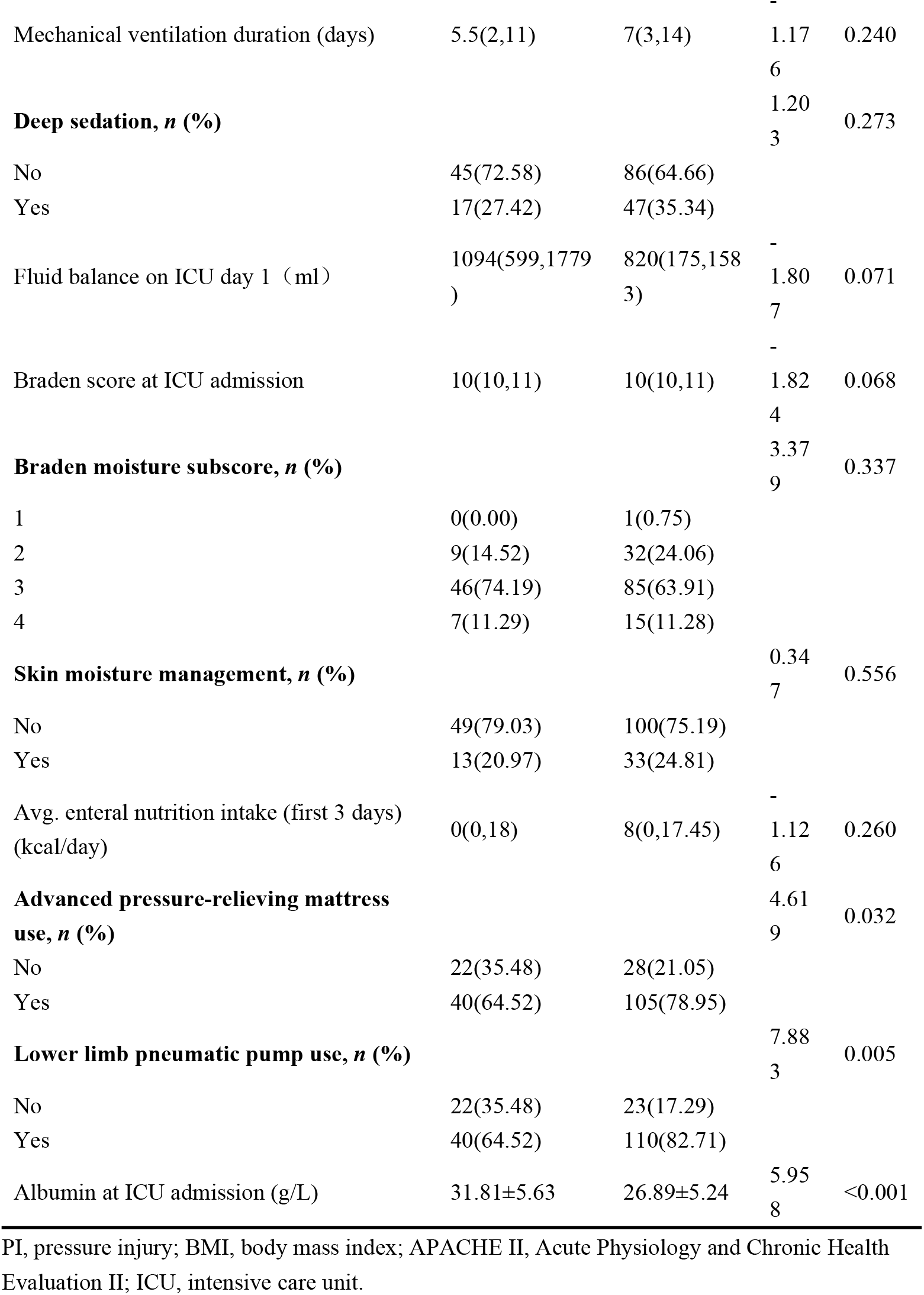
Comparison of baseline characteristics between non-severe and severe pressure injury groups in the training set.

### Predictor Selection and Model Construction

Variable selection was performed using Least Absolute Shrinkage and Selection Operator (LASSO) regression with 10-fold cross-validation. The optimal lambda value (λ = 0.0957) was determined as the lambda.1se, which yielded five predictors with non-zero coefficients: diabetes, maximum norepinephrine dose, use of a lower limb pneumatic pump, albumin level at ICU admission, and use of an advanced pressure-relieving mattress (Figure 2). These five variables were subsequently entered into the multivariable logistic regression model. As presented in Table 3, four of them were identified as independent predictors for severe pressure injuries: diabetes, maximum norepinephrine dose, use of a lower limb pneumatic pump, and albumin level at ICU admission.

**Table 3.**
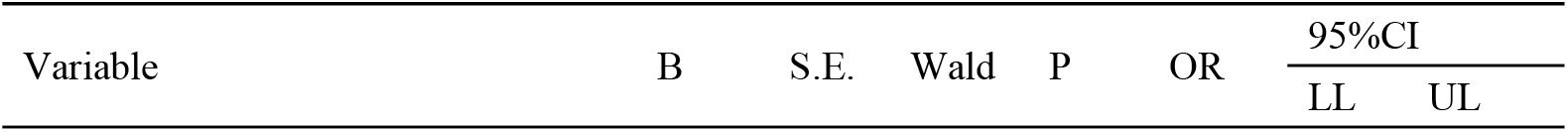
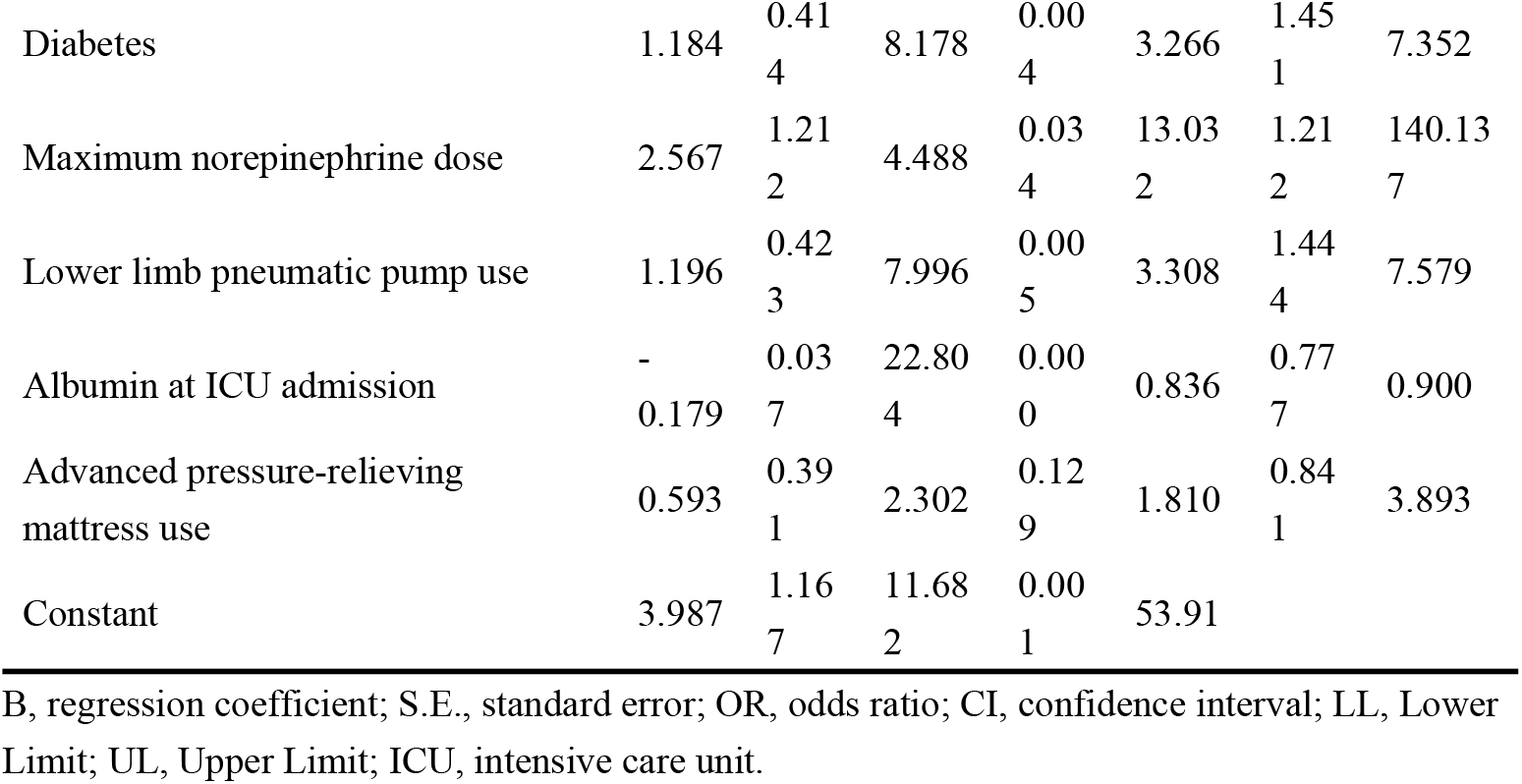
Multivariable logistic regression analysis for independent predictors of severe pressure injury.

| Variable | B | S.E. | Wald | P | OR | 95%CI |  |
| --- | --- | --- | --- | --- | --- | --- | --- |
|  |  |  |  |  |  | LL | UL |
| Diabetes | 1.184 | 0.41<br>4 | 8.178 | 0.00<br>4 | 3.266 | 1.45<br>1 | 7.352 |
| Maximum norepinephrine dose | 2.567 | 1.21<br>2 | 4.488 | 0.03<br>4 | 13.03<br>2 | 1.21<br>2 | 140.13<br>7 |
| Lower limb pneumatic pump use | 1.196 | 0.42<br>3 | 7.996 | 0.00<br>5 | 3.308 | 1.44<br>4 | 7.579 |
| Albumin at ICU admission | -<br>0.179 | 0.03<br>7 | 22.80<br>4 | 0.00<br>0 | 0.836 | 0.77<br>7 | 0.900 |
| Advanced pressure-relieving mattress use | 0.593 | 0.39<br>1 | 2.302 | 0.12<br>9 | 1.810 | 0.84<br>1 | 3.893 |
| Constant | 3.987 | 1.16<br>7 | 11.68<br>2 | 0.00<br>1 | 53.91 |  |  |

B, regression coefficient; S.E., standard error; OR, odds ratio; CI, confidence interval; LL, Lower Limit; UL, Upper Limit; ICU, intensive care unit.

**Figure 2.**
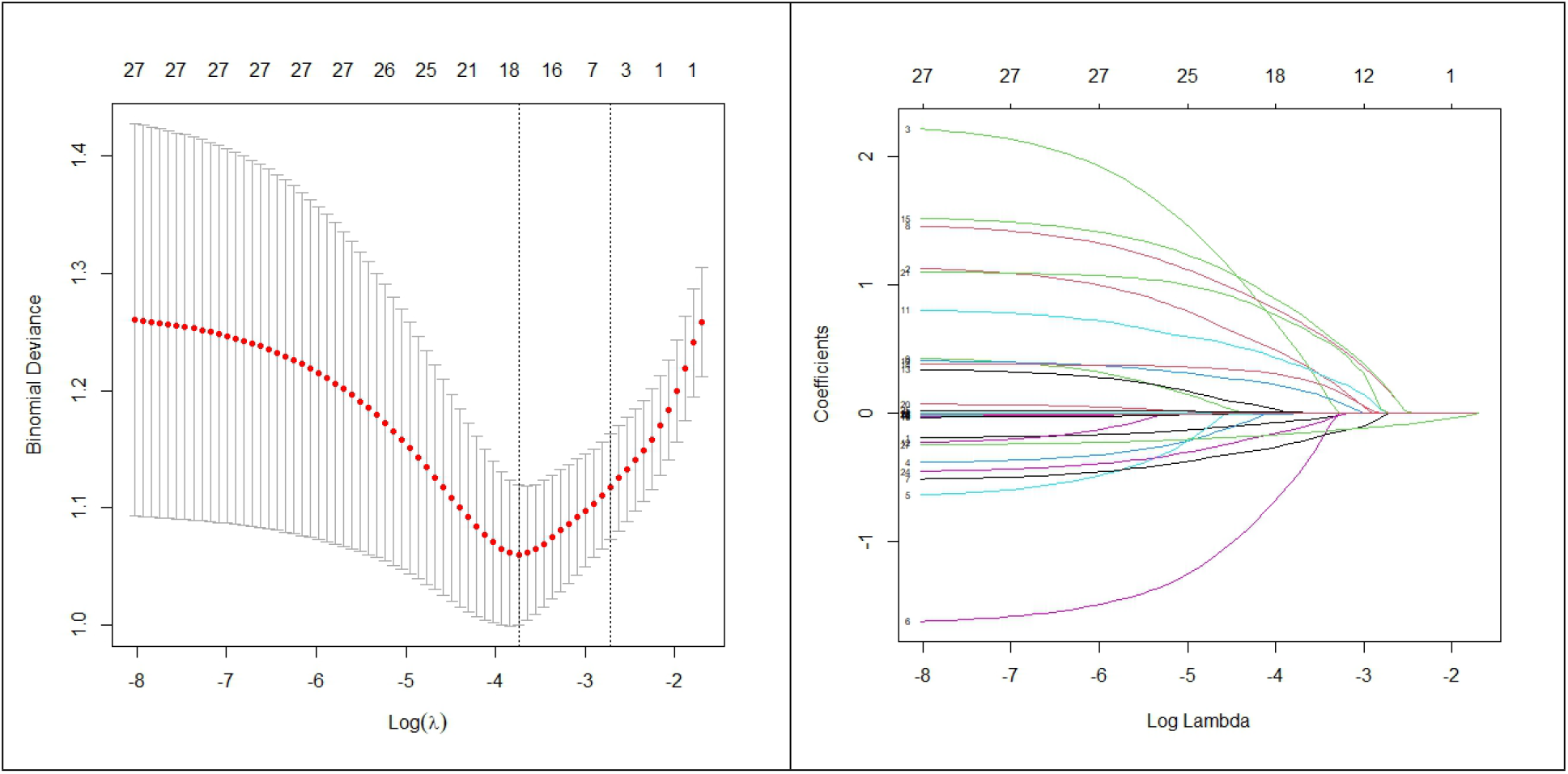
LASSO-logistic regression results. (A) Cross-validation plot. (B) Selection process by cross-validation method.

Based on the four independent predictors identified in the multivariable logistic regression analysis (Table 3) – a history of diabetes, maximum norepinephrine dose, use of a lower limb pneumatic pump, and serum albumin level at ICU admission – a nomogram was constructed to visualize the final prediction model (Figure 3). The nomogram provides a user-friendly tool for individualized risk estimation. Clinicians can obtain a patient’s total points by summing the individual scores assigned to each predictor variable. This total score corresponds to a specific predicted probability on the bottom scale, representing the individual’s risk of progressing from Stage 1 to severe (⩾ Stage 2) pressure injuries.

**Figure 3.**
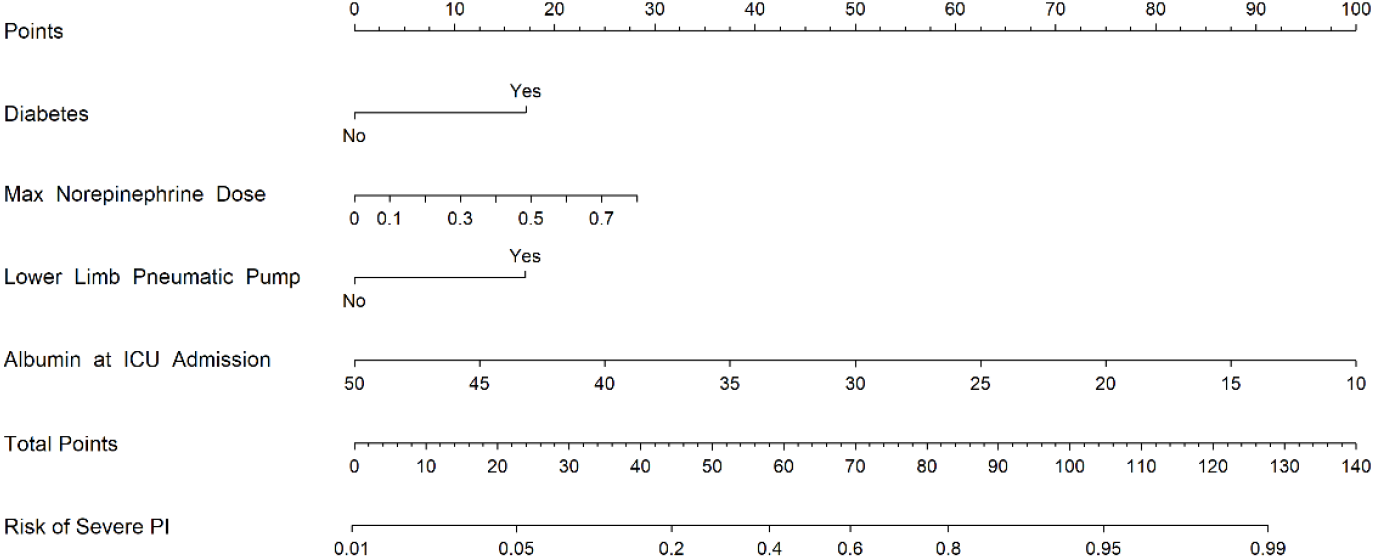
Nomogram for predicting the risk of severe pressure injury in ICU patients with Stage 1 injury. The nomogram incorporates four predictors: diabetes (yes/no), maximum norepinephrine dose (µg/kg/min), use of a lower limb pneumatic pump (yes/no), and serum albumin level at ICU admission (g/L). To use the nomogram, locate the patient’s value on each variable axis, draw a line upward to the “Points” axis to obtain the individual score, sum all scores to get the “Total Points”, and then draw a line downward from the “Total Points” axis to the bottom scale to read the predicted probability of severe pressure injury (PI) progression.

A nomogram was constructed based on these four independent predictors to visualize the final prediction model (Figure 3). This tool allows clinicians to estimate an individual patient’s risk of progressing from Stage 1 to severe (⩾ Stage 2) pressure injuries. The total points, obtained by summing scores from each predictor, correspond to a predicted probability on the bottom scale, facilitating rapid bedside risk assessment.

### Model Performance and Validation

The nomogram demonstrated good discriminatory ability, with an area under the curve (AUC) of 0.810 (95% CI: 0.748–0.872) in the training set and 0.805 (95% CI: 0.696–0.914) in the validation set (Figure 4). Calibration analysis revealed good model fit, indicated by Hosmer–Lemeshow test results (training set: χ^2^=4.535, *P*=0.806; validation set: χ^2^=7.987, *P*=0.435) and calibration curves showing close alignment between predicted and observed probabilities (Figure 5).

**Figure 4.**
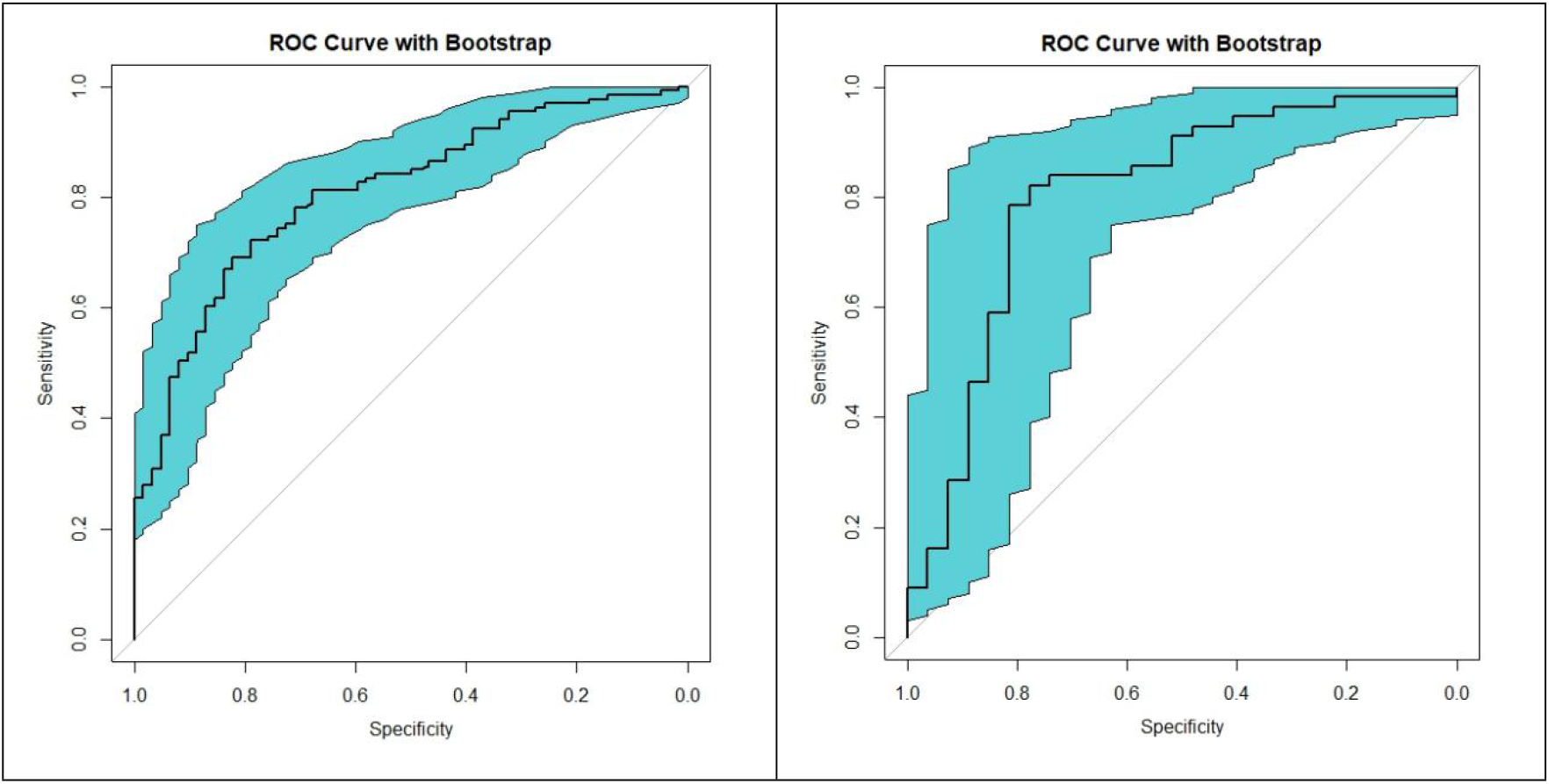
Bootstrap-enhanced receiver operating characteristic (ROC) curves of the nomogram for predicting severe pressure injury. (A) ROC curve in the training set. (B) ROC curve in the validation set.

**Figure 5.**
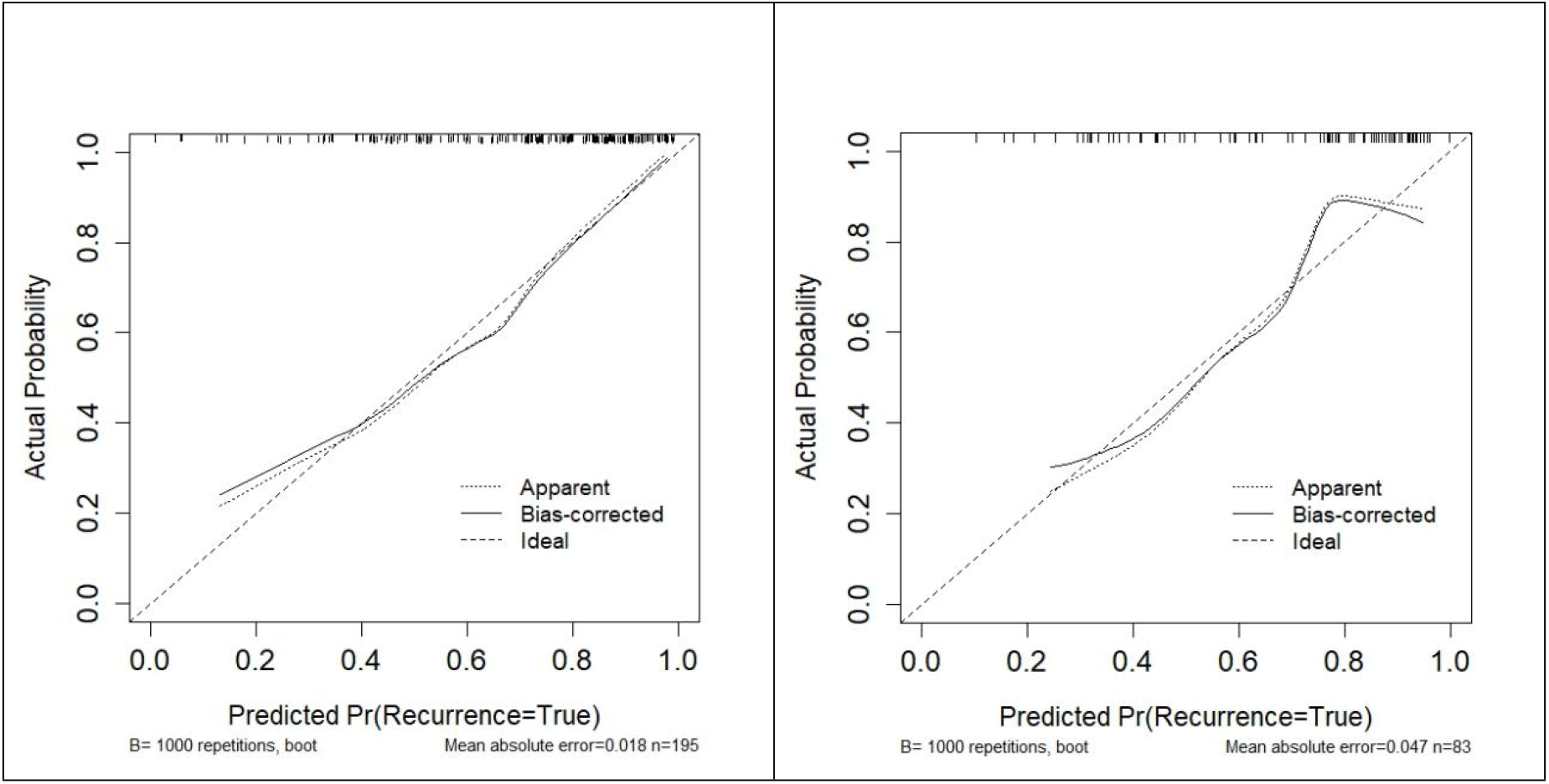
Calibration curves of the nomogram for predicting severe pressure injury. (A) Calibration curve in the training set. (B) Calibration curve in the validation set. B=1000 repetitions, boot, bootstrap repetitions.

Decision curve analysis (DCA) demonstrated that the nomogram provided a positive net benefit over a wide range of threshold probabilities in both the training and validation sets, supporting its clinical utility (Figure 6). Furthermore, the model showed robust performance and generalizability, as evidenced by a mean AUC of 0.799 (95% CI: 0.759–0.838) from ten-fold cross-validation (Figure 7).

**Figure 6.**
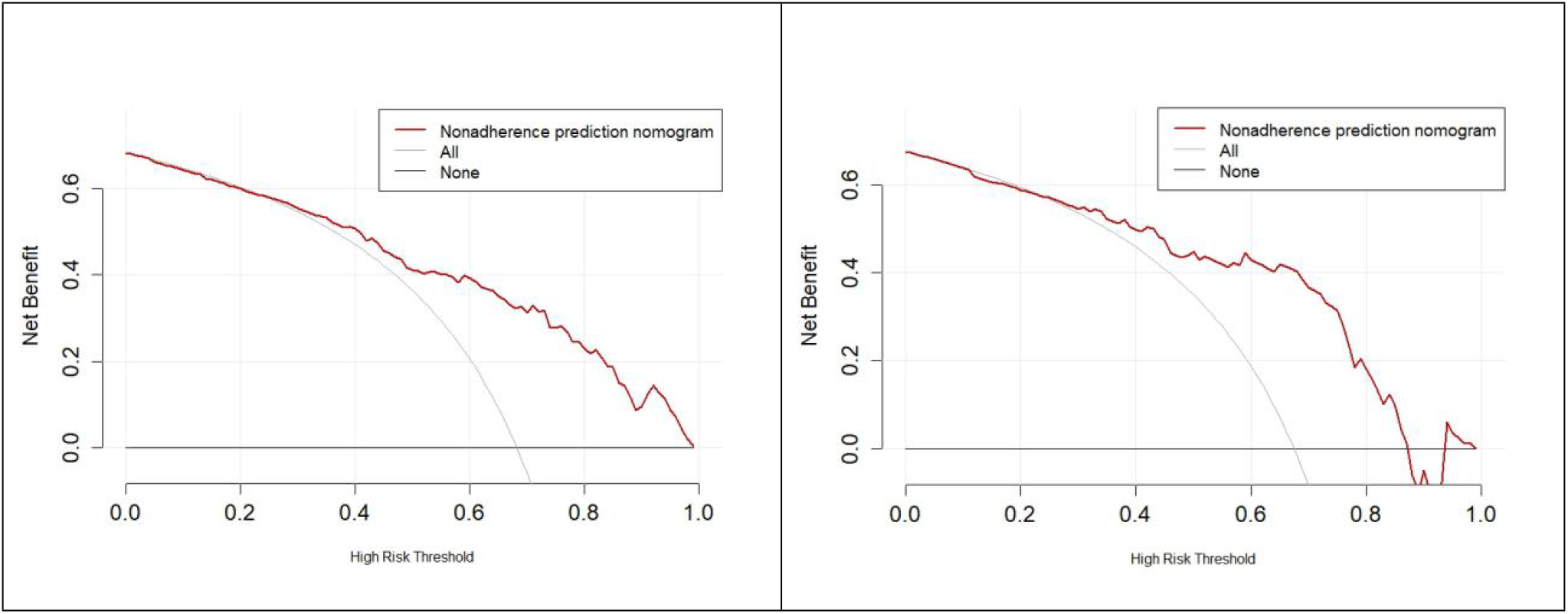
Decision curve analysis (DCA) of the nomogram for predicting severe pressure injury. (A) Net benefit curve in the training set. (B) Net benefit curve in the validation set.

**Figure 7.**
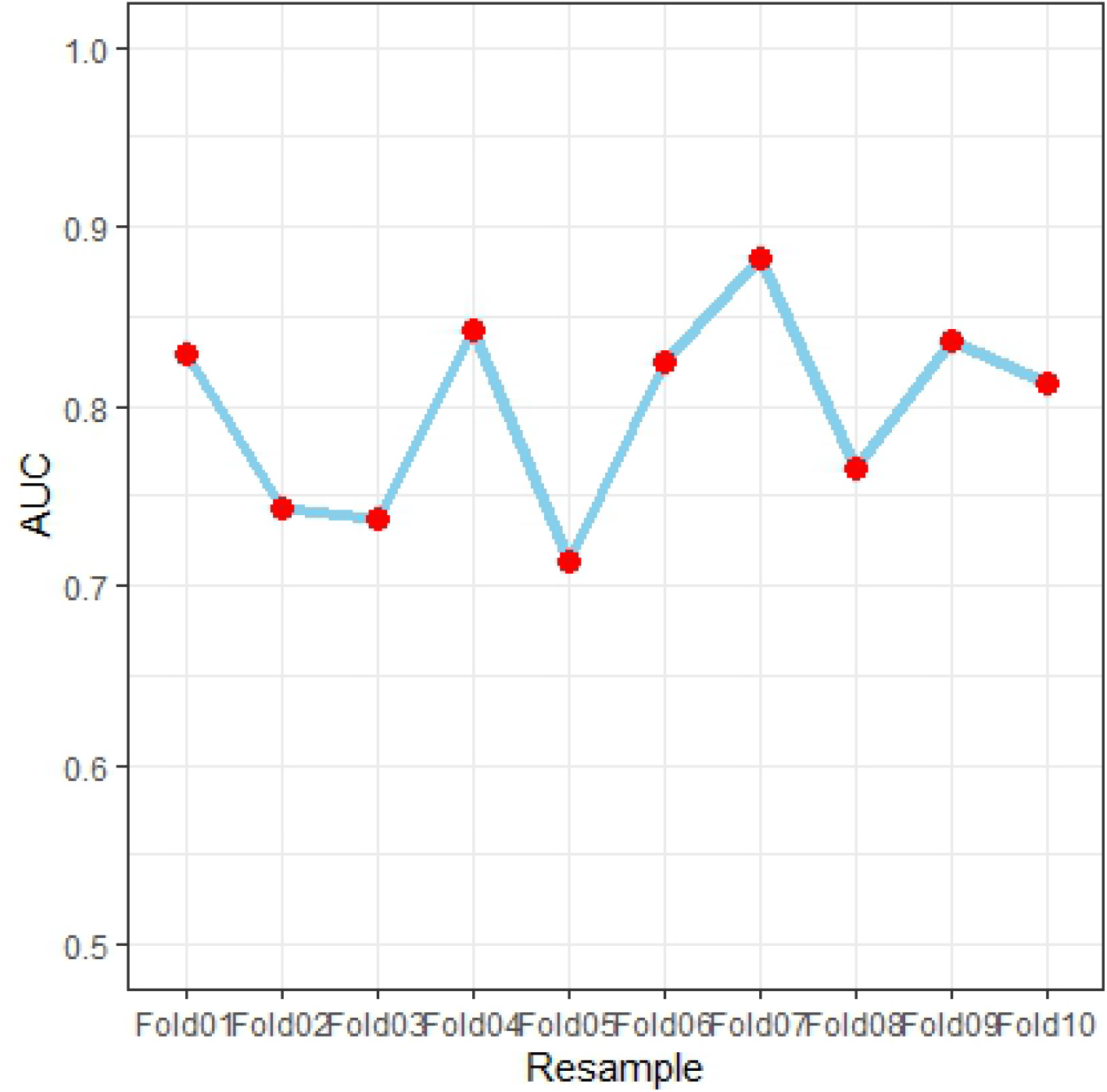
Performance of the nomogram for predicting severe pressure injury in ten-fold cross-validation. AUC, area under the curve.

### SHAP Analysis for Model Interpretation

SHapley Additive exPlanations (SHAP) analysis was performed to interpret the prediction model and quantify variable contributions. The SHAP feature importance plot (Figure 8A) identified albumin at ICU admission as the most influential predictor, followed by diabetes, maximum norepinephrine dose, and use of a lower limb pneumatic pump. The SHAP summary plot (Figure 8B) further illustrated the directional relationships: higher albumin levels consistently reduced predicted risk (negative SHAP values), while diabetes, elevated norepinephrine doses, and pneumatic pump use increased risk (positive SHAP values). These results align with the logistic regression findings and provide intuitive visual explanations of how each variable influences the prediction of severe pressure injury progression.

**Figure 8.**
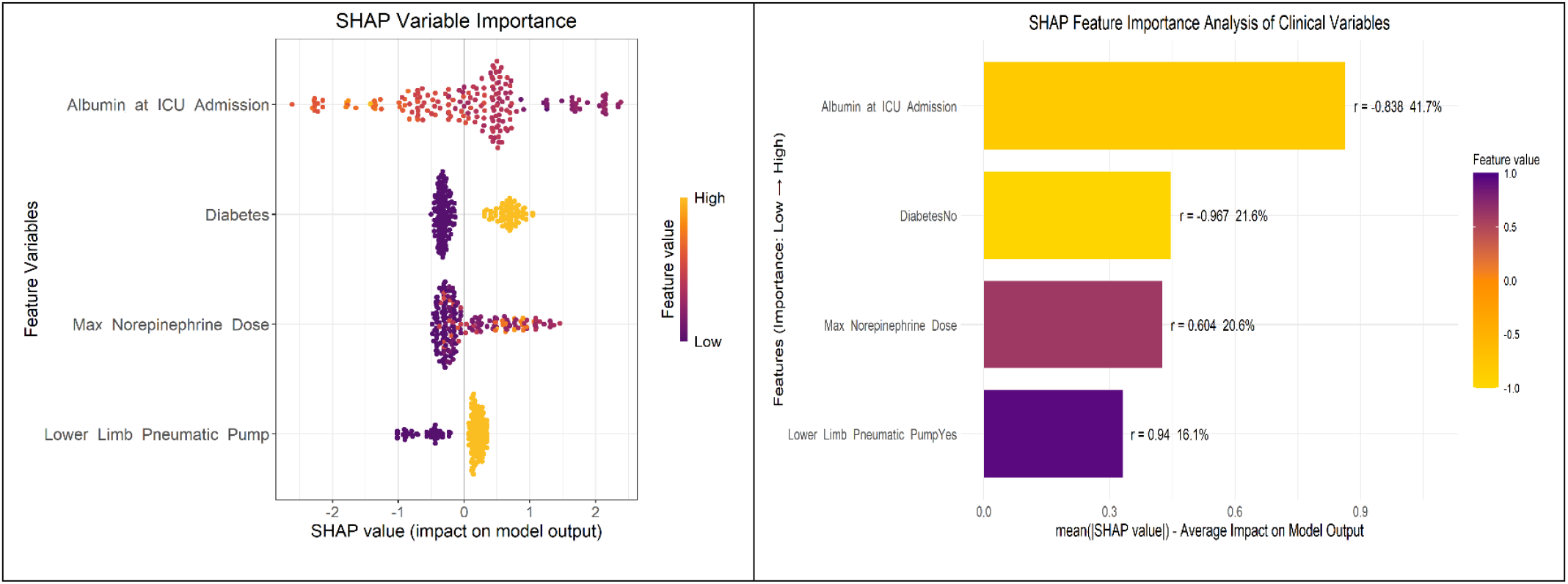
SHapley Additive exPlanations (SHAP) analysis for model interpretation. (A) SHAP feature importance plot. (B) SHAP summary plot.

### Subgroup Analysis

To evaluate the robustness and clinical applicability of the prediction model across different patient populations, we conducted comprehensive subgroup analyses based on septic shock status, mechanical ventilation requirement, and pressure injury location (Figures 9-11). These analyses revealed substantial heterogeneity in the effects of the predictors. In the septic shock subgroup, diabetes (OR=6.465, *P*<0.001) and maximum norepinephrine dose (OR=14.483, *P*=0.049) were significant risk factors, while albumin was protective (OR=0.863, *P*<0.001). Conversely, in non-septic shock patients, maximum norepinephrine dose (OR=39.219, *P*=0.046) and pneumatic pump use (OR=16.866, *P*<0.001) were strong risk factors, with albumin remaining protective (OR=0.767, *P*<0.001), while diabetes lost significance (*P*=0.185).

**Figure 9.**
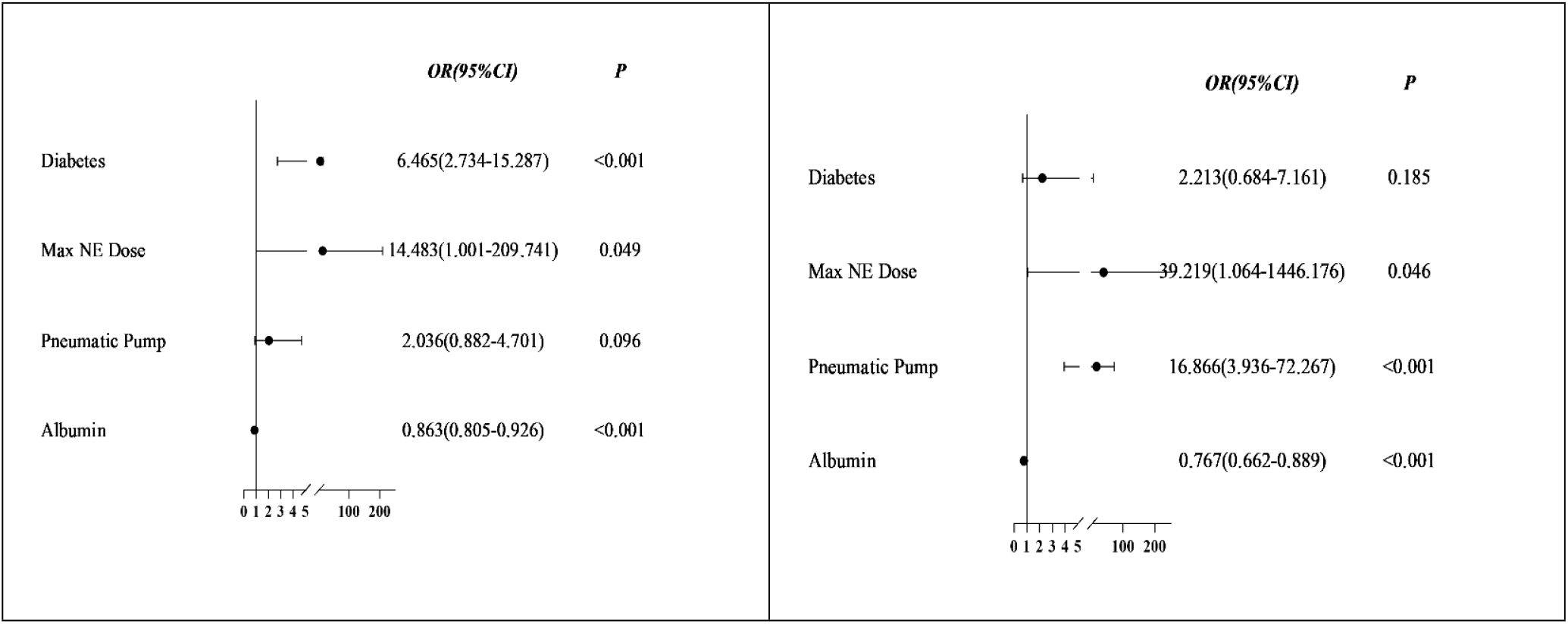
Forest plot of subgroup analysis based on septic shock status. (A) Septic shock subgroup. (B) Non-septic shock subgroup. Max NE Dose, maximum norepinephrine dose; Pneumatic Pump, lower limb pneumatic pump use.

**Figure 10.**
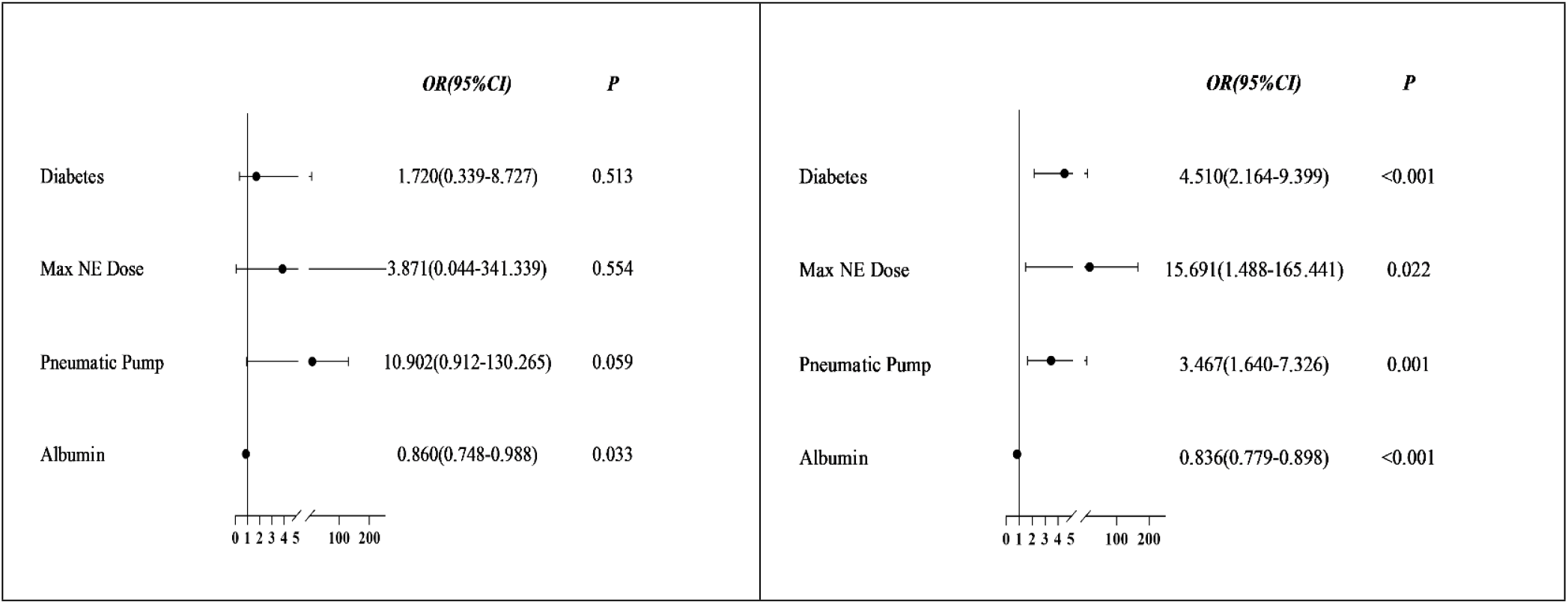
Forest plot of subgroup analysis based on mechanical ventilation status. (A) Non-mechanical ventilation (Non-MV) subgroup. (B) Mechanical ventilation (MV) subgroup. Max NE Dose, maximum norepinephrine dose; Pneumatic Pump, lower limb pneumatic pump use.

**Figure 11.**
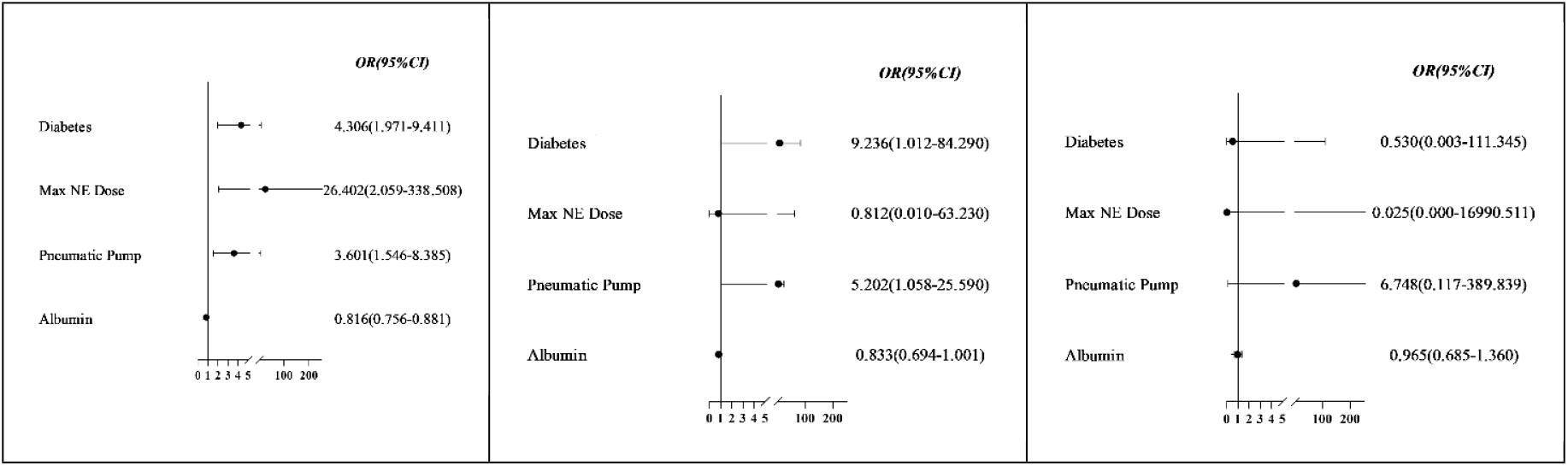
Forest plot of subgroup analysis based on the most severe pressure injury location. (A) Supine position group. (B) Lateral/positioning group. (C) Device-related group. Max NE Dose, maximum norepinephrine dose.

Stratification by mechanical ventilation status showed distinct patterns. Among non-ventilated patients, only albumin level was significantly protective (OR=0.860, *P*=0.033). In ventilated patients, diabetes (OR=4.510, *P*<0.001), maximum norepinephrine dose (OR=15.691, *P*=0.022), and pneumatic pump use (OR=3.467, *P*=0.001) were all significant risk factors, with albumin maintaining its protective effect (OR=0.836, *P*<0.001). Analysis by PI location revealed that all four predictors showed significant associations in the supine-position subgroup, while only diabetes and pneumatic pump use were significant for lateral/positioning-related injuries, and no significant associations were found in the device-related subgroup.

These stratified analyses confirm that while our core predictors demonstrate consistent directional effects, their magnitude and statistical significance vary substantially across clinical contexts. The model shows particularly strong predictive performance in septic shock patients and those requiring mechanical ventilation, while maintaining reliability across different injury locations, supporting its broad clinical applicability.

## Discussion

This study developed and validated a novel nomogram incorporating four readily available clinical factors (diabetes, maximum norepinephrine dose, use of a lower limb pneumatic pump, and serum albumin at ICU admission) to predict the progression from Stage 1 to severe PIs in critically ill patients. The model demonstrated good discrimination, calibration, and clinical utility, providing a practical tool for early risk stratification in this high-risk population.

The observed high incidence of severe PI progression in our cohort (68.2% in the training set) underscores the critical need for effective risk assessment in ICU patients presenting with Stage 1 PIs. Stage 1 PIs represent a pivotal point where timely intervention can prevent irreversible tissue damage. However, not all Stage 1 injuries progress, and current risk assessment tools, such as the Braden Scale, demonstrate limited accuracy in the dynamic ICU environment [12, 13]. This often leads to a generalized, inefficient allocation of preventive resources [14, 15]. Our model addresses this gap by enabling precise identification of the highest-risk individuals, allowing for targeted and intensified preventive strategies.

The independent predictors identified in our model are biologically plausible and reflect distinct pathophysiological pathways contributing to PI progression. **Hypoalbuminemia** was the strongest protective factor, a finding consistently linked to poor outcomes in critically ill patients [16]. Low serum albumin contributes to interstitial edema, impairs tissue repair, and serves as a marker of nutritional deficiency and systemic inflammation, all of which compromise skin integrity and wound healing [17, 18]. **Diabetes mellitus** is a well-established risk factor for PIs, primarily through its association with microvascular dysfunction, peripheral neuropathy, and impaired immune response, which collectively diminish tissue tolerance to pressure and ischemia [19, 20]. The **maximum norepinephrine dose** emerged as a potent risk factor, reflecting the severity of hemodynamic instability and vasopressor dependence. High-dose vasopressors, particularly norepinephrine, induce intense peripheral vasoconstriction, significantly reducing cutaneous blood flow and tissue perfusion, thereby predisposing patients to ischemic injury [21, 22]. Interestingly, the use of **lower limb pneumatic compression pumps** was associated with an increased risk, which appears counterintuitive as they are a preventive measure. This likely represents confounding by indication, where these devices are preferentially applied to patients who are perceived to be at the highest risk or are completely immobile, thus acting as a surrogate marker for profound immobility and critical illness severity rather than a causative factor [23, 24]. This highlights the importance of interpreting predictive factors within their clinical context. Our subgroup analyses revealed significant heterogeneity in predictor effects, enhancing the model’s clinical relevance. In septic shock patients, diabetes and vasopressor dose were dominant, emphasizing metabolic and perfusion-related risks [25, 26]. In contrast, among non-septic shock patients, immobility (proxied by pneumatic pump use) was a more prominent risk factor. For patients not requiring mechanical ventilation, albumin was the sole significant predictor, suggesting nutrition and baseline status are paramount in this less severely ill group. These findings advocate for a tailored approach to PI prevention based on the patient’s dominant clinical phenotype [27, 28].

The SHAP analysis provided an intuitive explanation of the model, confirming albumin as the most influential feature and clarifying the directional impact of each variable. This interpretability is crucial for fostering clinician trust and facilitating bedside application. Our model’s performance (AUC ∼0.81) is comparable to or better than previous prediction tools for hospital-acquired PIs in mixed populations, and it is specifically tailored for the critical transition from Stage 1 to more severe injury.

Several limitations should be acknowledged. First, the single-center, retrospective design may limit generalizability and introduce unmeasured confounding. Second, the use of a pneumatic pump as a variable is susceptible to indication bias. Third, external validation in independent, multi-center cohorts is necessary to confirm the model’s robustness and transportability. Future prospective studies should also evaluate the impact of implementing this nomogram on clinical outcomes and resource utilization.

## Conclusion

We developed a simple, interpretable nomogram that effectively predicts the risk of Stage 1 PI progression in ICU patients using four admission variables. By identifying high-risk individuals early, this tool can guide the rational allocation of intensive preventive measures, potentially mitigating the significant morbidity and healthcare costs associated with severe PIs.

## Data Availability

Data Availability Statement: The data underlying the results presented in this study are available from the Medical Ethics Committee of The Affiliated Lihuili Hospital of Ningbo University (contact via) for researchers who meet the criteria for access to confidential data. Data cannot be shared publicly because they contain sensitive patient information protected by privacy regulations and institutional data governance policies. All relevant aggregated data and statistical results are contained within the manuscript and its Supporting Information files. The analytic methods and code needed to reproduce the findings are available from the corresponding author upon reasonable request.

## Author contributions

Methodology: C.Z., W.W., H.X.

Investigation: C.Z.

Writing - original draft: C.Z.

Writing - review & editing: C.Z., W.W., H.X.

Supervision: H.X.

### Abbreviations

The following abbreviations are used in this manuscript:

APACHE II: Acute Physiology and Chronic Health Evaluation II
AUC: Area Under the Curve
CI: Confidence Interval
DCA: Decision Curve Analysis
ICU: Intensive Care Unit
LASSO: Least Absolute Shrinkage and Selection Operator
OR: Odds Ratio
PI: Pressure Injury
ROC: Receiver Operating Characteristic
SHAP: SHapley Additive exPlanations

## Notes

### Competing Interest Statement

The authors have declared no competing interest.

### Funding Statement

The author(s) received no specific funding for this work.

### Author Declarations

MedRxiv Ethics Details IRB/Oversight Body Name: Medical Ethics Committee of The Affiliated Lihuili Hospital of Ningbo University Approval Details: Approval Number: IIT2026SL0013 Approval Date: 12 January 2026 Approval Type: Full approval with waiver of informed consent Ethical Compliance Statement: This study was conducted in accordance with the Declaration of Helsinki. The need for informed consent was formally waived by the ethics committee due to the retrospective nature of the study. All patient data were anonymized and de-identified prior to analysis to protect confidentiality. Regulatory Classification: The research qualified for exemption from ongoing IRB oversight as it involved retrospective analysis of existing anonymized clinical data without patient intervention or identifiable information. Data Protection Measures: Data access restricted to authorized researchers All analytical outputs aggregated to prevent re-identification Secure storage on encrypted hospital servers with access logging Supplementary Materials Available: Full IRB approval documentation and waiver certification can be provided upon request to the corresponding author.

